# Urine-based multi-omic comparative analysis of COVID-19 and bacterial sepsis-induced ARDS

**DOI:** 10.1101/2022.08.10.22277939

**Authors:** Richa Batra, Rie Uni, Oleh M. Akchurin, Sergio Alvarez-Mulett, Luis G. Gómez-Escobar, Edwin Patino, Katherine L. Hoffman, Will Simmons, Kelsey Chetnik, Mustafa Buyukozkan, Elisa Benedetti, Karsten Suhre, Edward Schenck, Soo Jung Cho, Augustine M.K. Choi, Frank Schmidt, Mary E. Choi, Jan Krumsiek

## Abstract

Acute respiratory distress syndrome (ARDS), a life-threatening condition during critical illness, is a common complication of COVID-19. It can originate from various disease etiologies, including severe infections, major injury, or inhalation of irritants. ARDS poses substantial clinical challenges due to a lack of etiology-specific therapies, multisystem involvement, and heterogeneous, poor patient outcomes. A molecular comparison of ARDS groups holds the potential to reveal common and distinct mechanisms underlying ARDS pathogenesis. In this study, we performed a comparative analysis of urine-based metabolomics and proteomics profiles from COVID-19 ARDS patients (n = 42) and bacterial sepsis-induced ARDS patients (n = 17). The comparison of these ARDS etiologies identified 150 metabolites and 70 proteins that were differentially abundant between the two groups. Based on these findings, we interrogated the interplay of cell adhesion/extracellular matrix molecules, inflammation, and mitochondrial dysfunction in ARDS pathogenesis through a multi-omic network approach. Moreover, we identified a proteomic signature associated with mortality in COVID-19 ARDS patients, which contained several proteins that had previously been implicated in clinical manifestations frequently linked with ARDS pathogenesis. In summary, our results provide evidence for significant molecular differences in ARDS patients from different etiologies and a potential synergy of extracellular matrix molecules, inflammation, and mitochondrial dysfunction in ARDS pathogenesis. The proteomic mortality signature should be further investigated in future studies to develop prediction models for COVID-19 patient outcomes.

## 1. Introduction

The ongoing SARS-CoV-2 induced coronavirus disease 2019 (COVID-19) pandemic has been a major impediment to human life globally^1,2^. One of the main complications of severe COVID-19 is acute respiratory distress syndrome (ARDS). ARDS is a common presentation of critical illnesses, including severe infections, major injury, or inhalation of irritants^3^. While COVID-19-related ARDS and ARDS originating from other pathologies (hereby referred to as non-COVID-19 ARDS) have overlapping clinical features, COVID-19 ARDS is characterized by a protracted hyperinflammatory state and higher rates of thrombosis^4–13^. The field currently lacks etiology-specific therapies and reliable predictors of heterogeneous patient outcomes^14^.

To address these critical knowledge gaps, we recently elucidated molecular differences between and within two ARDS etiologies - COVID-19 and bacterial sepsis^15^. Extending this blood-based ARDS comparison, we here performed a similar analysis on urine samples. It has been suggested that urine-based molecular profiles reflect an individual’s physiological changes^16^ and have the potential to be used as diagnostic and prognostic biomarkers^17–20^. Previous urine-based COVID-19 studies have made substantial efforts to determine molecular markers distinguishing COVID-19 from healthy controls or less severe COVID-19 cases^21–24^. However, a detailed comparison of the molecular differences between two ARDS groups has so far been missing.

In this study, we analyzed urine samples from 59 ARDS patients, with COVID-19 (n = 42) and bacterial sepsis diagnosis (n = 17). We followed a two-step analysis workflow to elucidate the differences between the two ARDS groups. In the first part, we compared metabolomic and proteomic profiles between the two groups to identify differentially abundant molecules. For a systematic cross-omics analysis of these molecules, we performed a data-driven network analysis. In the second part of the study, we compared the molecular heterogeneity within each ARDS group. To this end, we associated the omics measurements with clinical manifestations, including acute kidney injury (AKI) incidence, platelet counts, PaO2/FiO2, and mortality. For further exploration and reproducibility of our findings, we share all results, analysis scripts, and de-identified omics data.

## 2. Results and Discussion

### 2.1. Molecular associations differentiating COVID-19 and bacterial sepsis-induced ARDS

To identify the molecular differences between COVID-19 ARDS and bacterial sepsis-induced ARDS, urine-based metabolomic and proteomic profiles from 59 samples were analyzed (n = 42 COVID-19, and n = 17 bacterial sepsis). At a 5% false discovery rate (FDR), 220 molecules were significantly different between the two groups, representing 150 metabolites (70 higher in COVID-19 ARDS and 80 lower), and 70 proteins (28 higher in COVID-19 and 42 lower) (**Figure 1a**). The results of this analysis are available in **Supplementary Table 1**. To aid the functional interpretation of these molecules, metabolites and proteins were annotated with ‘sub-pathway’ annotations provided by Metabolon and proteins were annotated with KEGG pathways^25^ (**Supplementary Table 2**). Top ranking pathways are shown in **Figure 1b**. Two of the pathways we identified in this ARDS comparison, extracellular matrix (ECM) and cell adhesion molecules (CAMs), have also been implicated in previous urine-based studies comparing COVID-19 with a control group^26^. In addition, blood-based studies have reported several of these pathways in the context of COVID-19 ARDS when compared to healthy controls, including amino acid metabolism, lipid metabolism, urea cycle, MAPK, PI3K-Akt, and JAK-STAT signaling^27–31^. Taken together, we identified 220 molecules that were differentially abundant between the two ARDS groups, with 33 distinct biological pathways that had three or more significant molecules.

**Figure 1:**
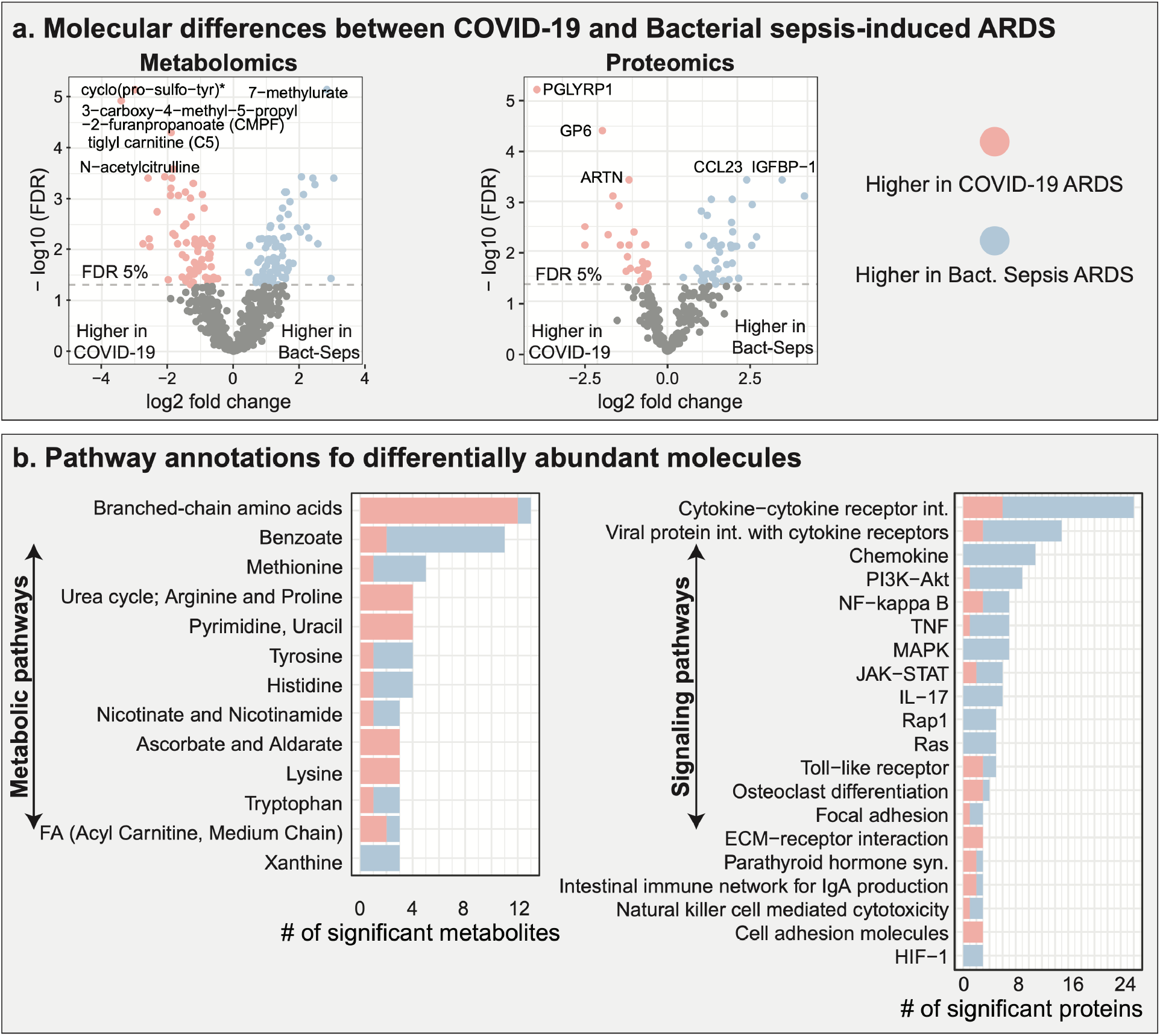
Molecular signature of COVID-19 ARDS compared to bacterial sepsis-induced ARDS. **a**. Differentially abundant molecules (150 metabolites, 70 proteins) between the two ARDS groups. **b**. Functional annotations of differentially abundant metabolites and proteins at the pathway level. Overall, 33 metabolic and signaling pathways with three or more significant molecules were deregulated between the two ARDS groups. FA=fatty acid.

### 2.2. ARDS-related interaction of mitochondrial dysfunction and ECM organization

Predefined pathway annotations provide context for already well-characterized biological processes; however, the insights they provide into cross-omics associations are limited. Therefore, we generated a data-driven multi-omic interaction network based on Gaussian graphical models (GGM)^32^. In earlier studies, we have shown that partial correlation-based GGMs reconstruct valid biochemical interactions from omics data in an unbiased fashion and can even identify previously unknown interactions between molecules^33–35^. The data-driven network contained 3,566 statistically significant interactions between the 708 metabolites and 266 proteins (**Figure 2a**). It was then annotated using the molecules that were differentially abundant between ARDS groups. An interactive version of the network is available in **Supplementary File 1** for further exploration.

**Figure 2:**
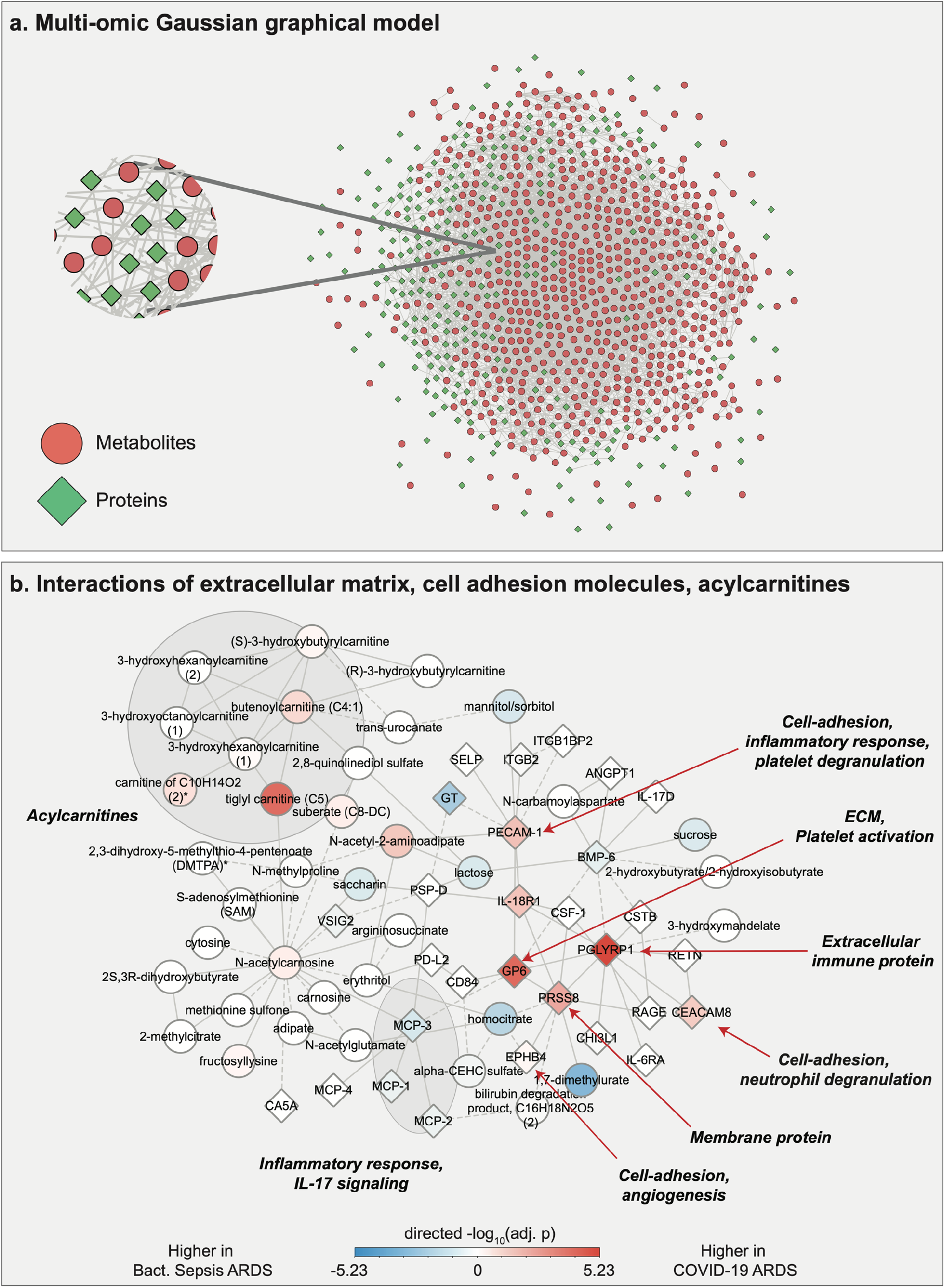
Multi-omic network and extracted ECM/CAM/acylcarnitine subnetwork. **a**. Gaussian graphical model (GGM) of metabolites and proteins. Shapes and colors of the molecules in the network are based on the two omics types. **b**. Subnetwork extracted from the full multi-omic GGM, built around tiglyl carnitine and GP6. The observed molecular interactions suggest an interplay of ECM derangement, inflammation, and mitochondrial dysfunction in ARDS pathogenesis.

We then generated a subnetwork focusing on several processes that have previously been implicated in COVID-19, namely mitochondrial dysfunction^36^, coagulopathy via cell-adhesion molecules (CAMs) and platelet activation^37,38^. To this end, we chose two molecules belonging to these processes which were also among the top metabolomic and proteomic hits in Figure 1a: Tiglyl carnitine, an acylcarnitine that represents mitochondrial function^39^, and glycoprotein 6 (GP6), which is involved in the extracellular matrix (ECM) and the platelet activation pathway.

The subnetwork was constructed by including tiglyl carnitine, GP6, and all of their first- and second-degree network neighbors, i.e., nodes that were separated from the two molecules by one or two edges in the network. The resulting subnetwork consisted of 66 molecules (37 metabolites, 29 proteins) with 106 interactions among them (**Figure 2b**). Within this subnetwork, tiglyl carnitine and GP6 were connected via MCP-3 and N-acetylcarnosine. The neighborhood of tiglyl carnitine consisted of other acylcarnitines, including butenoylcarnitine (C4:1), (S)-3-hydroxybutyrylcarnitine, and 3-hydroxyhexanoylcarnitine, all of which were higher in COVID-19 compared to bacterial sepsis-induced ARDS. The neighborhood of GP6 consisted of additional proteins related to ECM or CAMs, including EPHB4, CECAM8, and PECAM1, all of which were higher in COVID-19 compared to bacterial sepsis-induced ARDS. The mediating inflammatory MCP-3 protein was connected to other MCP proteins, which were higher in bacterial sepsis-induced ARDS than in COVID-19.

Overall, within the subnetwork, we observed cross-omics connections between clusters of CAMs/ECM and a group of acylcarnitines, mediated by a group of inflammatory MCP proteins. We speculate that the underlying interplay of these depicted biological processes might play a role in ARDS pathogenesis.

### 2.3. ARDS-specific heterogeneity of molecular associations across clinical manifestations

In the second part of our study, we tested ARDS group-specific molecular associations with four clinical manifestations: acute kidney injury (AKI), platelet count, patient’s oxygen in arterial blood to the fraction of the oxygen in the inspired air (PaO2/FIO2) ratio, and mortality. In the bacterial sepsis-induced ARDS group, no significant associations with any of these clinical manifestations were identified (5% FDR). In COVID-19 ARDS, there were 10 molecules associated with AKI, including 8 metabolites and 2 proteins, no molecules associated with platelet count, 6 metabolites associated with PaO2/FIO2, and 61 molecules associated with mortality, including 1 metabolite and 60 proteins. Thus, a molecular comparison of heterogeneous presentations across the two ARDS groups was not feasible. Detailed results are available in **Supplementary Tables 3 and 4**.

In the following, we focused on the proteomic mortality signature distinguishing survivors and non-survivors of COVID-19. Among 60 proteins that were significantly different between survivors and non-survivors, 22 were higher in survivors, 38 higher in non-survivors (**Figure 3a, left)**. Remarkably, in our recent plasma-based study^15^, we did not find any proteins that were associated with mortality in the same COVID-19 patients (**Figure 3a, right)**.

**Figure 3.**
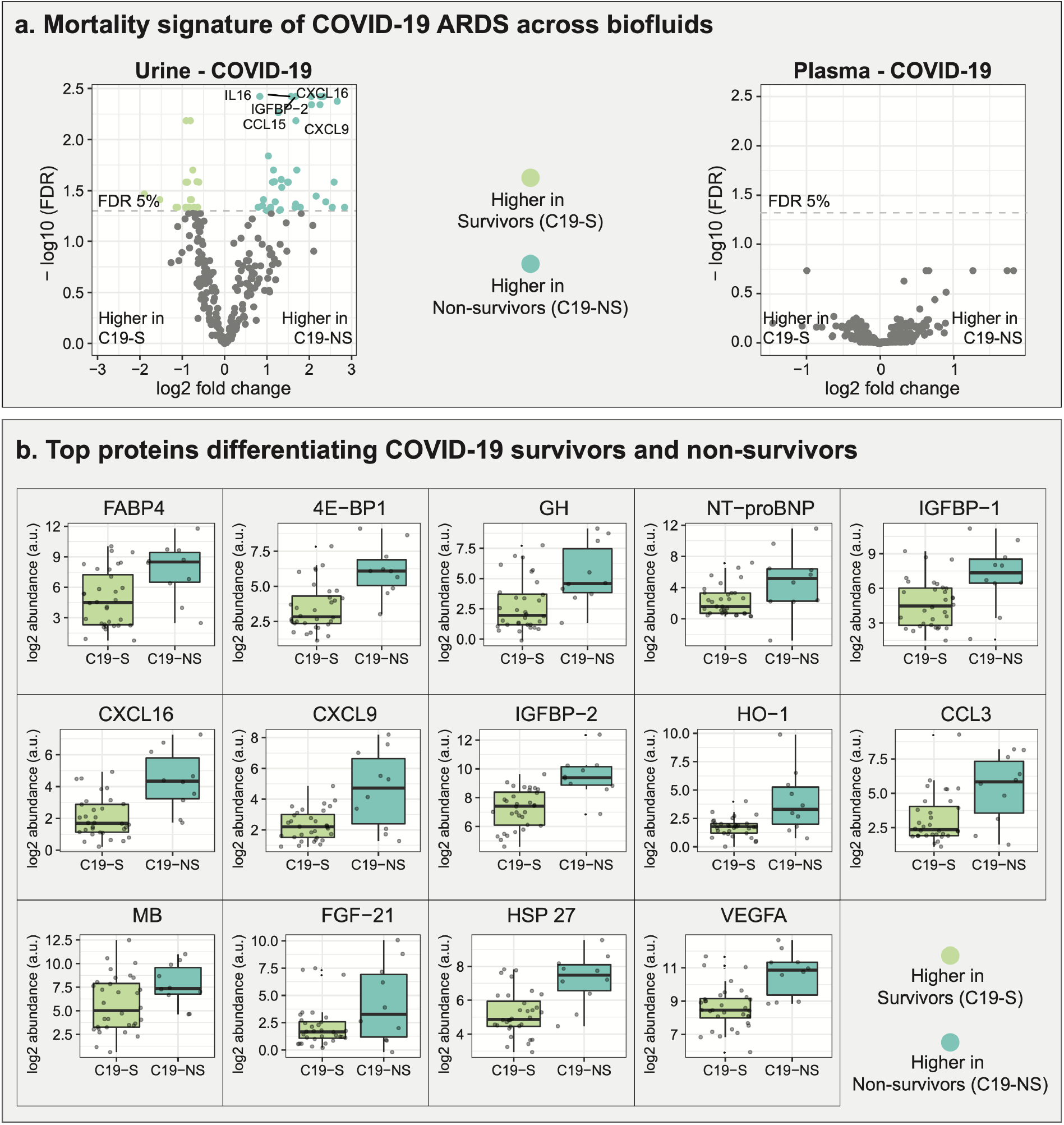
Proteomics-based mortality signature. **a**. Differentially abundant proteins in COVID-19 survivors and non-survivors, as observed in two bodily fluids, urine and plasma. 60 proteins were significant in urine proteomic profiles, while none of the proteins measured in the plasma of the same patients were associated with mortality. **b**. Top 14 differential proteins from COVID-19 urine-based mortality signature with log2 fold change larger than or equal to 2 at 5% FDR.

For further investigation of the urine-based COVID-19 mortality signature, we selected significant proteins with log2 fold changes larger than 2 (**Figure 3b**). Interestingly, several of these 14 proteins have previously been described as biomarkers of pathologies that are linked to ARDS. For example, NT-proBNP in the urine of preterm infants has been shown to inform about pulmonary hypertension^40^. IGFBP-2 is an indicator of pulmonary arterial hypertension (PAH)^41^ and can predict a decline of kidney function in type 2 diabetes^42^. FABP4 has been implicated in proteinuria and has been discussed as a marker of kidney glomerular damage^43^. CXCL16 is considered a urinary marker of poor renal outcome in diabetic kidney disease^44^. HO-1 is a candidate biomarker for oxidative damage in obstructive nephropathy^45^. VEGFA leads to increased inflammation in severe COVID-19^46^.

Taken together, these mortality-associated proteins have been implicated in ARDS-linked manifestations, including kidney dysfunction, pulmonary hypertension, and inflammation^8,47,48^. This provides insights into the potential pathophysiological processes behind the development of severe ARDS.

### 2.4. Conclusion

In this study, we presented a first urine-based multi-omic comparison of COVID-19 ARDS and non-COVID-19 ARDS. We compared 42 COVID-19 ARDS patients to 17 bacterial sepsis-induced ARDS patients using untargeted metabolomics (708 metabolites) and targeted proteomics (266 proteins). There were two main findings from our work. First, the multi-omic network approach highlighted the interplay of mitochondrial dysfunction and ECM derangement in ARDS pathogenesis. Second, we identified a proteomics-based mortality signature in COVID-19 ARDS patients. Notably, within the bacterial sepsis-induced ARDS group, no metabolites or proteins were found to be associated with any of the four clinical manifestations tested. In the following paragraphs, we discuss the two novel findings from our study.

Our multi-omic network-based analysis indicated an ARDS-related link between CAMs/ECM and mitochondrial dysfunction represented by acylcarnitines. In the analyzed subnetwork, the connections between these different biological processes were mediated by inflammatory proteins. Previous COVID-19 studies have already individually implicated these processes in ARDS, but have not proposed a link between these pathways in the context of ARDS^26,36,39^. Moreover, mechanistically, ECM, CAMs, and acylcarnitines have individually been linked with inflammation^49–51^. Our findings now highlight the potential synergy between these different cellular pathways in ARDS.

The proteomics-based mortality signature distinguishing COVID-19 survivors and non-survivors is another potentially novel finding from our study. Surprisingly, the mortality signal was absent in plasma proteomics profiles of the same patients. This could reflect frequent kidney involvement in severe COVID-19, which leads to the poor renal outcomes observed in our COVID-19 patients^48^. Moreover, the signature contains several proteins implicated in pathological processes that have been linked to ARDS, including inflammation, kidney dysfunction, and pulmonary hypertension^8,47,48^. In terms of clinical stratification approaches, a higher-powered study will be needed to assess whether machine learning models based on our signature are able to predict mortality in COVID-19 ARDS patients.

We recognize that our study design has several limitations. (1) Since the patients of the two ARDS groups were collected several years apart, differences in sample collection and storage protocols may lead to unaccountable variation across measurements. (2) Our cohort has a limited sample size (n = 59), with imbalanced ARDS groups (42 COVID-19 versus 17 bacterial sepsis patients). This relatively small sample size could have led to false negatives in our analysis, especially within the bacterial sepsis group. (3) Since the coverage of the metabolomics and proteomics platforms is limited, there is potential for missed associations with unmeasured molecules. (4) Our study was limited to statistical associations in a single cohort since we did not have access to an independent cohort for replication.

In conclusion, we presented a first urine-based multi-omic analysis of COVID-19 ARDS compared to bacterial sepsis-induced ARDS. Our analysis shows molecular similarities and differences between the two ARDS groups. The most striking finding was a proteomics-based mortality signature specifically for COVID-19 ARDS, which will require further investigation as a potential early biomarker for mortality.

### 3. Methods

### 3.1. Patient Population

The cohort was derived from the Weill Cornell Biobank of Critical Illness (WC-BOCI) at WCMC/NYP. The process for recruitment, data collection, and sample processing have been described previously^54–56^. In brief, the recruits in the WC-BOCI database were patients admitted to the intensive care unit with valid consent between October 2014 to May 2021, including 59 patients with COVID-19 ARDS (n=42) and bacterial sepsis-induced ARDS (n=17). Clinical data such as demographics, vital signs, labs, and ventilator parameters were obtained through the Weill Cornell-Critical Care Database for Advanced Research (WC-CEDAR) and the Weill Cornell Medicine COVID Institutional Data Repository (COVID-IDR). Additional clinical data were obtained through manual abstraction from the electronic health records.

This cohort included 47 (79.7%) males and 12 (20.3%) females, with a median age of 58.3. The overall mortality rate was 27.1 %, with 10 out of 42 in COVID-19 ARDS and 6 out of 17 in bacterial sepsis-induced ARDS. 45.8% of patients suffered from acute kidney injury (AKI), with 15 out of 42 in COVID-19 ARDS and 12 out of 17 in bacterial sepsis-induced ARDS. The sequential organ failure assessment (SOFA) index was comparable between the two groups, with a median of 10 in the COVID-19 group and 9 in the bacterial sepsis group. Detailed demographics of the patient cohort are provided in **Supplementary Table 5**.

### 3.2. Clinical manifestations

Below are the definitions used to diagnose the clinical manifestations used in this study.

#### Acute Respiratory Distress Syndrome (ARDS)

ARDS was assessed using the Berlin definition^57^, and followed by a review of the subject’s history, arterial blood gas, and chest X-ray by two independent pulmonary and critical care attendings to adjudicate the diagnosis. For bacterial sepsis-induced ARDS, an additional criterion was used as outlined in The Third International Consensus Definitions for Sepsis and Septic Shock^58^. For diagnosis of COVID-19, a positive viral swab of the nasopharynx tested for SARS-CoV-2 via RT-PCR was required.

#### *Acute Kidney Injury (AKI)*. ‘Kidney Disease

Improving Global Outcomes’ definition (KDIGO) was used to diagnose AKI. To this end, either of the following criteria was required: (a) serum creatinine change of greater than or equal to 0.3 mg/dL within 48 hours, (b) serum creatinine greater than or equal to 1.5 times the baseline serum creatinine known or assumed to have occurred within the past 7 days, (c) urine output less than or equal to 0.5 mL/kg/hour for six hours^59^.

### 3.3. Sample handling

Urine specimens were obtained from patients admitted to ICU at WCMC/NYP. Briefly, urine samples were centrifuged, and the supernatant was stored at -80°C until the omics profiling was performed. An electronic informed consent was obtained from all subjects for inclusion.

### 3.4. Proteomic profiling

Proteomic profiling was performed by the Proteomics Core of Weill Cornell Medicine-Qatar using the Olink platform (Uppsala, Sweden)^15^. Briefly, manufacturer’s instructions were followed to profile the samples using four panels including Inflammation, Cardiovascular II, and Cardiovascular III panels. Thorough quality assurance/quality control (QA/QC) was performed to monitor the assay’s incubation, extension, and detection steps. For (Ct) value extraction, Fluidigm’s reverse transcription-polymerase chain reaction (RT-PCR) analysis software was used at a quality threshold of 0.5 and linear baseline correction. Further processing of Ct values was performed using the Olink NPX manager software (Olink, Uppsala, Sweden).

### 3.5. Metabolomic profiling

Metabolic profiling was performed by Metabolon, Inc (Morrisville, NC) using ultrahigh performance liquid chromatograph-tandem mass spectroscopy (UPLC-MS/MS)^15^. Briefly, samples were subjected to methanol extraction and then divided into four aliquots for each of the mass spectroscopic methods. Rigorous quality assurance/quality control (QA/QC) was performed to monitor instrument performance and aid in chromatographic alignment. The four mass spectroscopic methods used were optimized for acidic positive ion hydrophilic compounds, acidic positive ion hydrophobic compounds, and basic negative ions, the fourth aliquot was analyzed via negative ionization. For metabolite identification, Metabolon’s proprietary software was used to deliver high-quality abundances of metabolites.

### 3.6. Data processing

Metabolomic and proteomic profiles were preprocessed before downstream analysis: Molecules with more than 25% missing values were removed, leaving 708 out of 1,112 metabolites and 266 out of 276 proteins. Probabilistic quotient normalization^60^ was used to correct sample-wise variation in the data. Data was log_2_ transformed, followed by k-nearest-neighbor-based imputation^61^ for the remaining missing values. Abundance levels of the following ten proteins were measured in duplicates by Olink panels and were therefore averaged: CCL3, CXCL1, FGF-21, FGF-23, IL-18, IL-6, MCP-1, OPG, SCF, and uPA. All data processing was performed using the maplet R package^62^.

### 3.7. Differential analysis of molecules

For association analysis, we used linear models with metabolites/proteins as the dependent variable and diagnosis/clinical manifestations as independent variables. Further factors such as age, sex, and BMI were not used as covariates in the models, since they are considered determinants of disease severity themselves^63^. To control the false discovery rate, the Benjamini-Hochberg (BH) method^64^ was used to correct p-values. All analyses were performed using the maplet R package^62^.

### 3.8. Pathway annotation and filtering

For functional annotation of the differently abundant molecules, we used Metabolon’s ‘sub-pathway’ groups and signaling pathways from KEGG^25^ for metabolites and proteins, respectively. **Supplementary Table 2** contains the complete list of annotations. For our analysis, we considered Metabolon’s sub-pathways with the term ‘metabolism’ and non-disease KEGG pathways with at least 3 significant molecules.

### 3.9. Multi-Omic network inference

To generate a multi-omic data-driven network we created a Gaussian graphical model (GGM) using the GeneNet R package^32^. GGMs are a partial correlation-based approach for identifying statistical connections among the molecules. To construct the network, pair of molecules (nodes) with significant partial correlations at 5% FDR were included and were connected with an edge. Following this, these nodes were annotated based on the statistical association results between the ARDS groups. To this end, a *p*_*score*_ was computed using the following formula: *p*_*score*_ = − log_10_(*p*.*adj*). *d*,, where *p. adj* is the adjusted p-value of the association, and *d* is the direction (-1/1) of the association based on test statistic (positive or negative association with the outcome). This score was used to color the nodes in the network.

## Data Availability

All data produced are available online at

https://doi.org/10.6084/m9.figshare.20260998.v1

## Study Approval

The study was approved by the institutional review board at Weill Cornell Medicine (protocol number 22-03024534). Written informed consent was received before participation by all patients, except when the institutional review board approved a waiver of informed consent (e.g., for the use of discarded samples and de-identified patient data).

## Data and code availability

The preprocessed data used in this study can be downloaded at https://doi.org/10.6084/m9.figshare.20260998.v1

All R scripts to generate the tables and figures of this paper are available at https://github.com/krumsieklab/covid-ards-urine

## Author contributions

RB, RU, AMKC, OMA, MEC, and JK designed the study. SJC and ES contributed to study design discussions. KS and FS contributed to proteomics profiling. SAM, LGGE, RU, WS, and KLH extracted de-identified clinical data. LGGE and EP collected samples. EB, MB, and KC contributed to statistical discussions. RB performed the analysis. RB, RU, OMA, MEC, and JK interpreted the results. RB, RU, MEC, and JK wrote the manuscript. MEC and JK supervised the study. All authors approved the final manuscript.

